# A Pilot Study of an Esophageal Cooling Device During Radiofrequency Ablation for Atrial Fibrillation

**DOI:** 10.1101/2020.01.27.20019026

**Authors:** Brad Clark, Nazia Alvi, Joseph Hanks, Brad Suprenant

**Author notes:** **Corresponding author:** Brad Clark, DO, St. Vincent Hospital, 8333 Naab Rd #400, Indianapolis, IN, 46260, United States of America. **Author contributions:** BC: concept, design, execution, data collection, data analysis/interpretation, manuscript drafting; NA: data analysis/interpretation; JH: design, data analysis/interpretation; BS: concept, design, data analysis/interpretation; all authors provided critical revision and approval of the manuscript. **Conflict of interest:** None.

## Abstract

**Background:** Esophageal thermal injury is a risk of ablation of the posterior left atrium despite various devices utilized to date.

**Objective:** Evaluate the potential of a commercially-available esophageal cooling device to provide esophageal protection during left atrial catheter ablation.

**Methods:** In this pilot study, we randomized 6 patients undergoing catheter ablation for atrial fibrillation. Three patients received standard of care for our site (use of a single-sensor temperature probe, with adjunct iced-water instillation for any temperature increases >1°C). Three patients received standard ablation after placement of the esophageal cooling device using a circulating water temperature of 4°C. All patients underwent transesophageal echocardiogram (TEE) and esophagogastroduodenoscopy (EGD) on the day prior to ablation followed by EGD on the day after.

**Results:** In the 3 control patients, one had no evidence of esophageal mucosal damage, one had diffuse sloughing of the esophageal mucosa and multiple ulcerations, and one had a superficial ulcer with large clot. Both patients with lesions were classified as Zargar 2a. In the 3 patients treated with the cooling device, one had no evidence of esophageal mucosal damage, one had esophageal erythema (Zargar 1), and one had a solitary Zargar 2a lesion. At 3-month follow-up, 1 patient in each group had recurrence of atrial fibrillation.

**Conclusions:** The extent of esophageal injury was less severe with a commercially available esophageal cooling device than with reactive instillation of ice-cold water. This pilot study supports further evaluation with a larger clinical trial.

## Introduction

The treatment of atrial fibrillation via pulmonary vein isolation using radiofrequency (RF) ablation has well documented success. A rare complication of this procedure is atrial-esophageal fistula (AEF) formation.[1,2] A precursor to AEF involves esophageal submucosal and possibly mucosal injury.[3] This damage is delivered by heating of the posterior aspect of the left atrium to create a transmural infarction that may inadvertently cause further thermal injury to esophageal tissue. Esophageal mucosal injury occurs in up to 30-50% of patients undergoing ablation.[4]

Attempts to protect the esophagus have been evaluated including mechanical displacement of the esophagus as well as multiple cooling modalities. Cooling approaches that have been evaluated include ice lavage, a cooling sac (EPSac, RossHart Technologies Inc.), and multiple types of cooling balloons. Many of these devices have been tested in bench models and animals with limited human use.[5-12] However, these earlier attempts at esophageal protection have not completely eliminated the risk of esophageal damage.

One commonly used approach to esophageal protection involves luminal esophageal temperature (LET) monitoring with adjustment in ablation location and timing based on temperature change. However, the efficacy of this approach remains uncertain.[13] Moreover, it has been suggested that the LET probe also contributes to thermal injury.[14] For this reason, we performed a pilot study of a commercially available esophageal cooling device currently utilized for whole-body temperature management, and shown to have protective effects in animal and mathematical models, as well as in recently presented clinical data.[15,16] The primary objective was to evaluate the potential to utilize this device for the reduction in the occurrence and severity of esophageal injury in patients undergoing RF ablation.

## Methods

### Study population

Six adults diagnosed with an atrial arrhythmia were included in this blinded (patient and outcome assessor), randomized controlled pilot study. Patients considered for ablation of the posterior wall (non-pregnant, between age 18-90 years, and without prior esophageal damage or disease) were asked to participate in the study after written informed consent. The study was approved by the hospital’s institutional review board, and the study was registered on Clinicaltrials.gov with identifier #NCT03481023.

Permutated block randomization was used for group designation. The study included 5 male and 1 female subjects aged 55 to 71 years. Two of the patients had mild reduction in left ventricular ejection fraction (LVEF) with the remainder of the patients having normal LVEF. Pre-procedural diagnoses included paroxysmal atrial fibrillation (5 of 6), typical atrial flutter (1 of 6), and atypical atrial flutter (2 of 6). All patients had failed at least 1 antiarrhythmic agent prior to the procedure. An outpatient visit was arranged at 3 months following ablation to assess for adverse events and to arrange further evaluation of atrial arrhythmia recurrence. (Table 1)

**Table 1.**
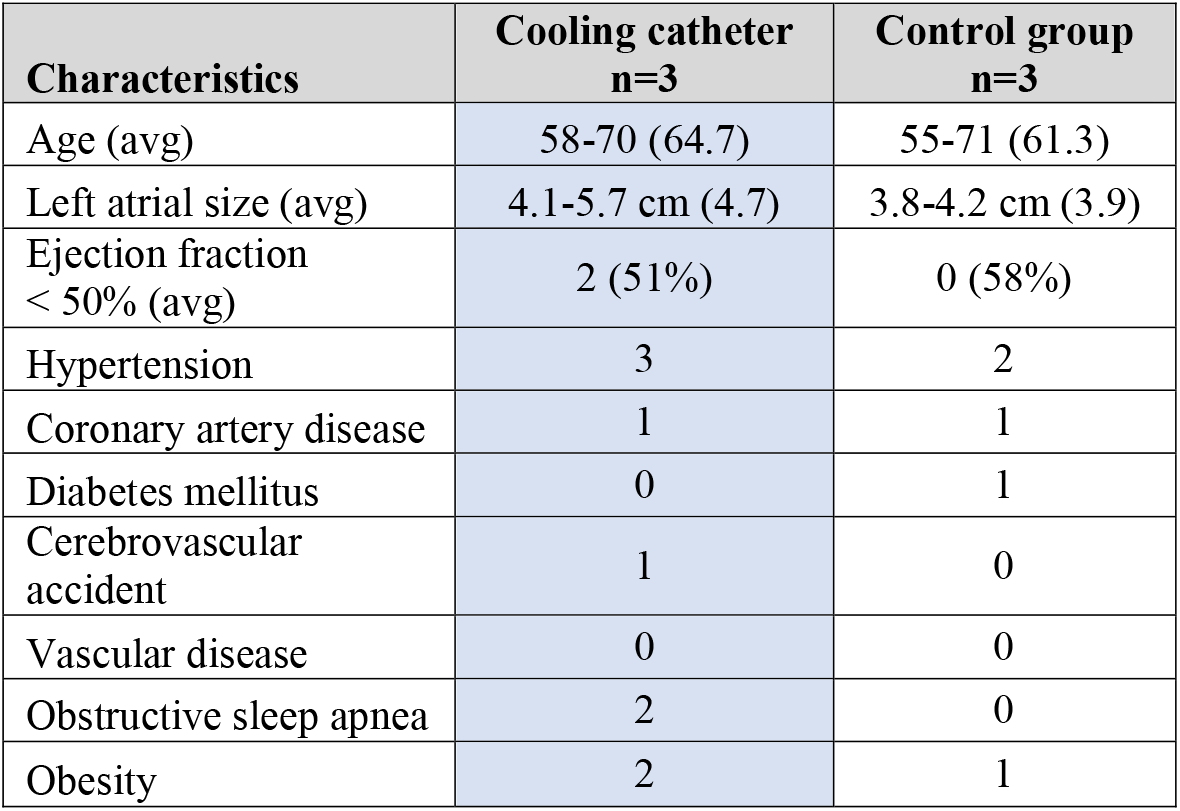
Patient characteristics

### Intervention

Patients received a transesophageal echocardiogram (TEE) the day prior to ablation, which was coupled with endoscopic evaluation including esophagogastroduodenoscopy (EGD). An independent gastroenterologist blinded to the patient’s assignment graded esophageal mucosal damage. We used Zargar’s Modified Endoscopic Classification Scheme to provide objective quantification of mucosal changes (Table 2). This classification scheme expands on the customary endoscopic classification of burns (grades 0 to 3) by subdividing grade 2 burns into 2a and 2b, and grade 3 burns into 3a and 3b for prognostic and therapeutic implications.

**Table 2.** Zargar classification and its corresponding endoscopic description

| **Zargar classification** | **Description** |
| --- | --- |
| Grade 0 | Normal mucosa |
| Grade I | Edema and erythema of the mucosa |
| Grade IIA | Hemorrhage, erosions, blisters, superficial ulcers |
| Grade IIB | Circumferential lesions |
| Grade IIIA | Focal deep gray or brownish-black ulcers |
| Grade IIIB | Extensive deep gray or brownish-black ulcers |
| Grade IV | Perforation |

All patients had general anesthesia as is customary for this institution. All ablations were performed by the same electrophysiologist with the choice of ablation including any combination of pulmonary vein isolation, true left atrial roof line, mitral line, cavo-tricuspid isthmus, and complex fractionated atrial electrogram ablation as was clinically indicated. Patients received a single-sensor temperature probe placed in the esophagus, as well as a standard nasogastric tube placed to the depth of the left atrium. The temperature readings were monitored by lab staff during ablations on the posterior wall. In the event that temperature rose by more than 1°C from baseline, ablation would cease, and the anesthesiologist was instructed to instill ice-cold water into the nasogastric tube in 10-20 mL aliquots. Ablation would then resume after equilibration of temperature readings to patient baseline. Ablation was carried out with 35 watts of energy at 55 degree temperature on the posterior wall and 50 watts of energy and 55 degree temperature on the anterior wall using a 4mm tip RF catheter. The RF catheters were non-contact-force sensing and non-irrigated.

The treatment group included the same pre-operative assessment with TEE followed by EGD on the day prior to ablation, and post-operative care and assessment was also identical to the control group. The treatment group patients received placement of a commercially available esophageal cooling device (figure 1) into the esophagus. This device provides a closed-circuit flow of water through a multi-channel cylindrical silicone tube placed in the esophagus analogously to a standard orogastric tube, heating or cooling a patient through conductive heat transfer across the esophagus, and convective heat transfer through the device.[17] The device was connected to a heat exchange unit (Blanketrol III Hyper-Hypothermia System, Gentherm Medical, Cincinnati, OH) which allows for large volume flow of temperature regulated distilled water at a rate of 136 L/hour. Just prior to ablation the coolant was set to 4°C, and this temperature was maintained during ablation on the posterior wall. The patient’s core temperature was monitored via a rectal temperature probe.

**Fig. 1.**
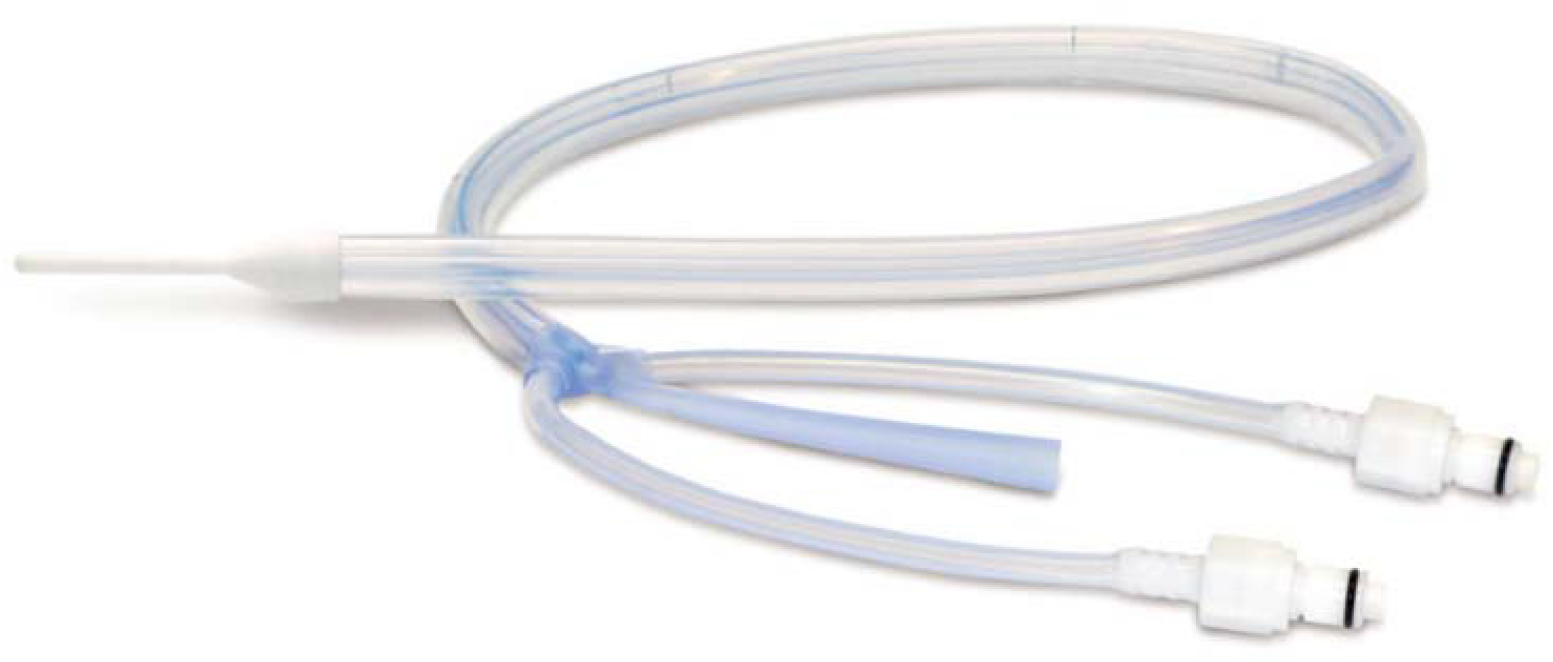
EnsoETM esophageal cooling catheter. Manufactured by Attune Medical, Chicago, IL

All patients were observed in the hospital overnight with repeat EGD performed on the morning following the ablation procedure. Follow-up care included a 1-2 week assessment as well as a 3 month visit. All patients had an event monitor or loop recorder between 3-6 months following ablation.

## Results

All of the patients had normal esophageal mucosa immediately following TEE on the day prior to ablation. One patient was incidentally noted to have Barret’s esophagus which required long-term monitoring. In the 3 control patients, all had multiple instillations of ice-cold water for temperature excursions above 1 C during posterior wall ablation. Of these 3 patients, one had no evidence of esophageal mucosal damage, one had diffuse sloughing of the esophageal mucosa and multiple ulcerations, and one had a superficial ulcer with large clot. Example EGD images of control patient lesions are shown in Figure 2. Both patients with lesions were classified as Zargar 2a (Table 3).

**Table 3.** Study results. Results of all patients including indication for procedure, esophageal findings, and post-op monitoring. The total number of ablation lesions and the number of lesions applied to the posterior wall are also included for comparison between patients. AF = atrial fibrillation; CFAE = complex fractionated atrial electrogram; CTI = cavotricuspid isthmus; ECD = Esophageal cooling device; LA = left atrium.

| Study patient | Age/<br>Gender | ECD | Indication | Zagars<br>Pre-op/<br>Post-op | Post-op<br>ulcers | Procedure | Total<br>ablation<br>lesions | Posterior<br>wall<br>lesions | Post-op monitor |
| --- | --- | --- | --- | --- | --- | --- | --- | --- | --- |
| 186-1 | 58 F | No | Paroxysmal AF | 0/0 | 0 | PVI with LA roof line | 273 | 45 | No AF on loop recorder |
| 186-2 | 58 M | Yes | Paroxysmal AF, and typical atrial flutter | 0/2a | 1 | PVI, roof line, and CTI line | 128 | 21 | Palpitations - AF noted on event recorder at 30 days |
| 186-3 | 66 M | Yes | Paroxysmal AF, and atypical atrial flutter | 0/0 | 0 | PVI, roof line, mitral isthmus line, CTI and CFAE ablation | 511 | 118 | Palpitations - No AF or aflutter on event recorder |
| 186-4 | 55 M | No | Paroxysmal AF | 0/2a | Multiple | PVI | 320 | 48 | No AF on event monitor at 6 months |
| 186-5 | 70 M | Yes | Atypical atrial flutter | 0/1 | 0 ulcers - Erythema noted | Roof line, mitral isthmus line, CTI line, and CFAE ablation | 273 | 5 | Palpitations (much improved) - No AF on Event Monitor |
| 186-6 | 71 M | No | Paroxysmal AF | 0/2a | 1 | PVI, roof line | 511 | 118 | AF noted on loop recorder at 30 days |

**Fig. 2.**
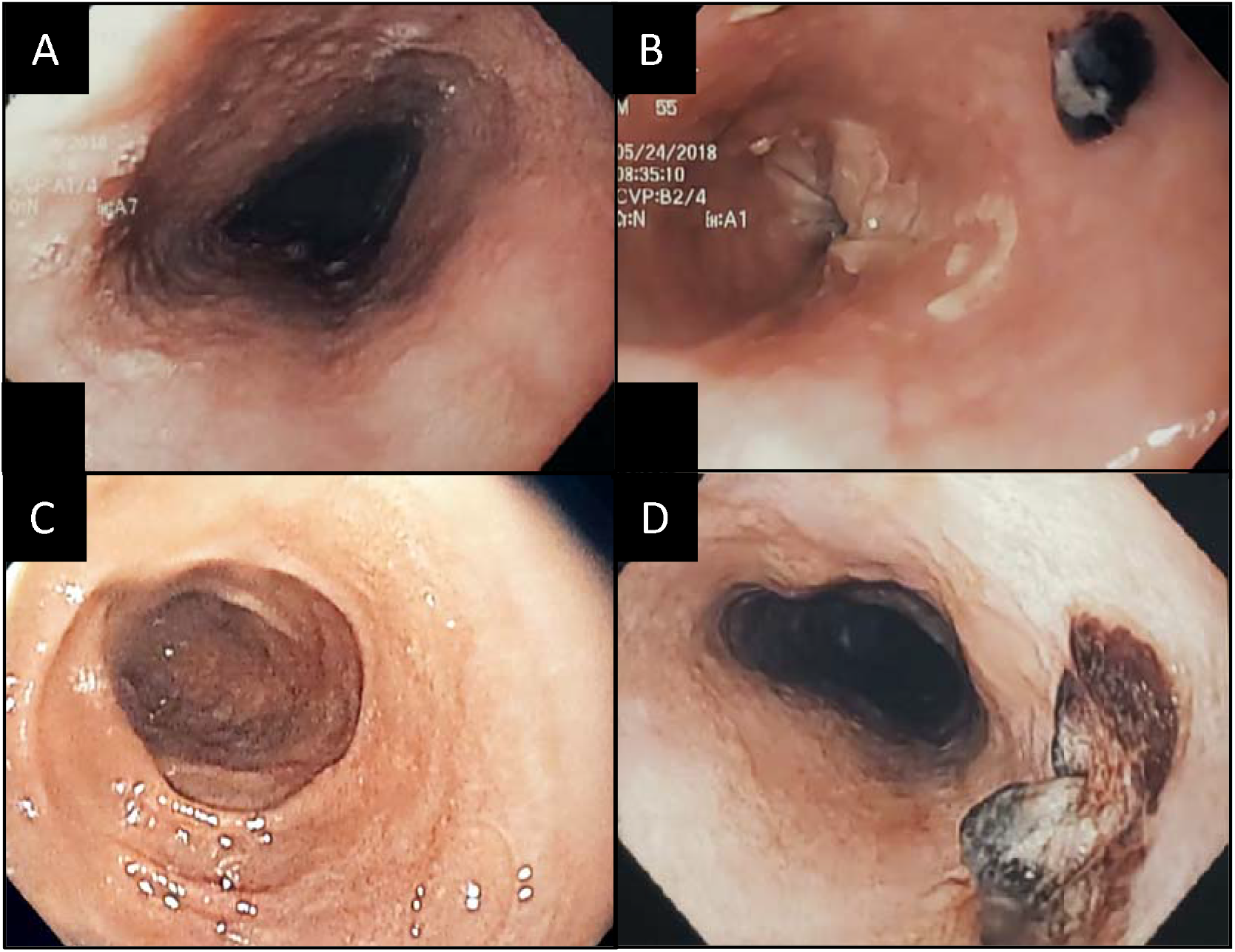
EGD images in control patients showing esophageal ulceration. Image A shows a patient with normal mucosa pre-ablation, and image B shows multiple ulcers with Zargars Grade 2a damage post-ablation, with diffuse sloughing as well as multiple ulcerations, with the largest shown in the image. Image C shows another control patient with normal esophageal mucosa pre-ablation, and image D shows a large ulceration with necrotic core in the same patient post-ablation

In the 3 patients treated with the esophageal cooling device, one had no evidence of esophageal mucosal damage, one had esophageal erythema (Zargar 1), and one had a solitary Zargar 2a lesion. There was no significant difference in the total ablation time or posterior wall ablation between the groups, as seen in Table 4. It is notable that the patient with the most extensive posterior wall ablation in the treatment group did not have any esophageal injury (Table 3). Example EGD images of the treatment arm patients are shown in Figure 3. Placement of the esophageal cooling device was straightforward and did not interfere with ablation, allowing the procedure to continue without interruption for temperature overshoot. Moreover, there was no need for fluoroscopy use since the cooling device stays placed to the depth of the stomach without need for repositioning during the procedure.

**Table 4.** Ablation data averaged for the control group and the cooling catheter group. Note that despite variability in ablation time between patients there is no significant difference between the groups. The Mann-Whitney U test was used to obtain significance. RF = radio frequency.

|  | Esophageal Cooling Device Used |  |  |  | P-value |
| --- | --- | --- | --- | --- | --- |
|  | No |  | Yes |  |  |
|  | Mean | Standard Deviation | Mean | Standard Deviation |  |
| Total Ablation Time (seconds) | 3700 | 958 | 3432 | 1385 | 1.0 |
| Total number of RF applications | 368 | 126 | 304 | 193 | 0.7 |
| Posterior wall lesions | 70 | 41 | 48 | 61 | 0.4 |
| Average power (W) | 34 | 9 | 31 | 11 | 1.0 |
| Maximum power (W) | 40 | 8 | 44 | 22 | 0.7 |
| Average impedance (Ohms) | 59 | 3 | 76 | 11 | 0.1 |
| Average temp (C) | 46 | 2 | 47 | 4 | 1.0 |
| Maximum temp (C) | 56 | 4 | 60 | 8 | 0.7 |

**Fig. 3.**
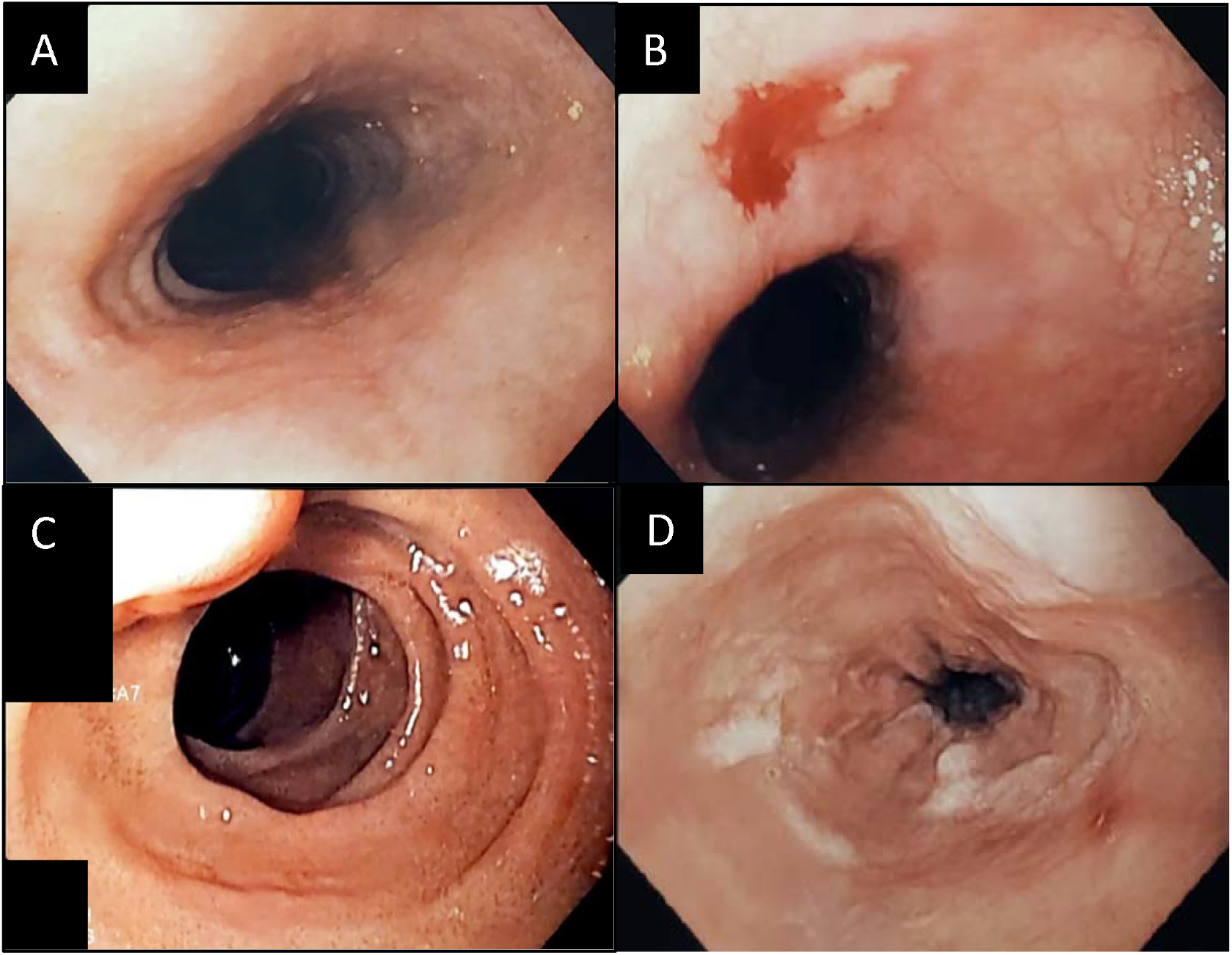
EGD images in cooling catheter patients showing esophageal changes. Image A shows a patient with normal mucosa pre-ablation and image B shows an ulcer with post-ablation superficial ulcer (Zargars grade 2a). Image C shows a patient with normal mucosa pre-ablation and image D shows post-ablation erythema (Zargars grade 1)

The core temperature of all subjects was monitored via rectal temperature sensor. There was minimal systemic cooling effect noted. The device was additionally used as an aid in rewarming the patient at the conclusion of the procedure by setting the system to a warming mode. As commonly described, the systemic temperature of the control patients was also noted to reduce with general anesthesia despite the use of forced-air blanket warming.

Patients were evaluated via event monitor or loop recorder. Evaluation at between 3-6 months after the ablation procedure showed recurrence of atrial fibrillation in one control patient and one treatment arm patient.

## Discussion

In this first randomized, controlled pilot study of a commercially available esophageal cooling device utilized for esophageal protection during left atrial ablation for the treatment of atrial fibrillation, we found fewer high-grade lesions compared to control patients receiving intermittent instillation of ice-cold water. Specifically, the treatment arm cohort included a single ulcer in one patient and an erythematous patch in another patient. The control group included two patients with esophageal lesions with a higher degree of injury, diffuse sloughing, and larger ulcerations.

Although not powered for statistical significance, the potential reduction of lesion incidence and severity may offer a new option for esophageal protection and the prevention of AEF. In addition, because stopping ablation for temperature rises in the esophagus did not occur, and because repositioning of the traditionally-used temperature probe was not required, an improvement in workflow, with a corresponding reduction in fluoroscopy usage, is suggested.

Various approaches have been developed to attempt esophageal protection during RF ablation, including LET monitoring, cooling, and deviating the esophagus. Nevertheless, currently available discrete sensor probes, whether single or multiple, do not appear to significantly reduce injury rates, and there is a potential for esophageal harm with esophageal deviation [18-20] In contrast, multiple earlier studies have evaluated esophageal cooling using a variety of techniques, with all but one suggesting potential benefit.[5-12] The availability now of a commercial device currently on the market for whole-body temperature modulation offers the chance to further study this modality, which in our pilot data, appears to offer potential. A number of larger clinical studies of this approach are ongoing: Improving Oesophageal Protection During AF Ablation (IMPACT) - NCT03819946, Esophageal Cooling for AF Ablation (eCoolAF) - NCT03691571, and Esophageal Damage Protection During Pulmonary Vein Ablation. Pilot Study. - NCT03832959.

## Limitations

We were not able to blind the physician performing the ablation, and as such, differences in ablation technique may have occurred. Nevertheless, because cessation of ablation occurred regularly in the control arm after elevated temperature probe readings, but did not occur in the treatment arm, it is likely that the actual regional energy deposition over time was greater in the treatment arm. Our standard practice also utilizes non-contact-force sensing, non-irrigated catheters. However, non-irrigated and non-contact-force sensing catheters are used by 30% of the writing group of the 2017 HRS-EHRA-ECAS-APHRS-SOLAECE Expert Consensus Statement on Catheter and Surgical Ablation of Atrial Fibrillation, so generalizability is not precluded.[21] We attempted to include assessment of the submucosal tissue architecture using endoscopic ultrasound (EUS) to identify tissue abnormalities which may be missed on EGD; however, technical challenges rendered this uninformative. These technical challenges included difficulties in determining exact longitudinal distance of the ultrasound probe relative to EGD scope, and uncertainties in determining abnormal findings, since EUS has not commonly been used for this particular purpose.

## Conclusions

The extent of esophageal injury was less severe with a commercially-available esophageal cooling device than with periodic manual instillation of ice-cold water, and there was no reduction in ablation efficacy. Moreover, the efficiency of ablation procedures was improved by eliminating stoppage for temperature overshoot or temperature probe repositioning. This pilot study supports further evaluation with a larger clinical trial.

## Data Availability

Available data will be provided in the manuscript without external links.

